# Multiclass arrhythmia classification using multimodal smartwatch photoplethysmography signals collected in real-life settings

**DOI:** 10.1101/2024.12.03.24318445

**Authors:** Dong Han, Jihye Moon, Luís Roberto Mercado Díaz, Darren Chen, Devan Williams, Fahimeh Mohagheghian, Om Ghetia, Andrew G. Peitzsch, Youngsun Kong, Nishat Nishita, Ohm Ghutadaria, Taylor A. Orwig, Edith Mensah Otabil, Kamran Noorishirazi, Alexander Hamel, Emily L. Dickson, Danielle DiMezza, Darleen Lessard, Ziyue Wang, Jordy Mehawej, Andreas Filippaios, Syed Naeem, Matthew F. Gottbrecht, Timothy P. Fitzgibbons, Jane S. Saczynski, Bruce Barton, Eric Y. Ding, Khanh-Van Tran, David D. McManus, Ki H. Chon

## Abstract

In the early stages of atrial fibrillation (AF), most cases are paroxysmal (pAF), making identification only possible with continuous and prolonged monitoring. With the advent of wearables, smartwatches equipped with photoplethysmographic (PPG) sensors are an ideal approach for continuous monitoring of pAF. There have been numerous studies demonstrating successful capture of pAF events, especially using deep learning. However, deep learning requires a large amount of data and independent testing on diverse datasets, to ensure the generalizability of the model, and most prior studies did not meet these requirements. Moreover, most prior studies using wearable-based PPG sensor data collection were limited either to controlled environments, to minimize motion artifacts, or to short duration data collection. Most importantly, frequent premature atrial and ventricular contractions (PAC/PVC) can confound most AF detection algorithms. This has not been well studied, largely due to limited datasets containing these rhythms. Note that the recent deep learning models show 97% AF detection accuracy, and the sensitivity of the current state-of-the-art technique for PAC/PVC detection is only 75% on minimally motion artifact corrupted PPG data. Our study aims to address the above limitations using a recently completed NIH-funded Pulsewatch clinical trial which collected smartwatch PPG data over two weeks from 106 subjects. For our approach, we used multi-modal data which included 1D PPG, accelerometer, and heart rate data. We used a computationally efficient 1D bi-directional Gated Recurrent Unit (1D-Bi-GRU) deep learning model to detect three classes: normal sinus rhythm, AF, and PAC/PVC. Our proposed 1D-Bi-GRU model’s performance was compared with two other deep learning models that have reported some of the highest performance metrics, in prior work. For three-arrhythmia-classification, testing data for all deep learning models consisted of using independent data and subjects from the training data, and further evaluations were performed using two independent datasets that were not part of the training dataset. Our multimodal model achieved an unprecedented 83% sensitivity for PAC/PVC detection while maintaining a high accuracy of 97.31% for AF detection. Our model was computationally more efficient (14 times more efficient and 2.7 times faster) and outperformed the best state-of-the-art model by 20.81% for PAC/PVC sensitivity and 2.55% for AF accuracy. We also tested our models on two independent PPG datasets collected with a different smartwatch and a fingertip PPG sensor. Our three-arrhythmia-classification results show high macro-averaged area under the receiver operating characteristic curve values of 96.22%, and 94.17% for two independent datasets, demonstrating better generalizability of the proposed model.

## Introduction

Atrial fibrillation (AF) is the most common serious cardiac dysrhythmia, and the incidence and prevalence of AF are increasing worldwide^1^. Long-term monitoring for AF is usually effective for incident AF detection, even though most early cases of AF are brief, asymptomatic, and intermittent^2^ (hence known as paroxysmal AF (pAF)). While detection of pAF requires long term monitoring, the electrocardiogram (ECG) devices developed for long term monitoring have poor patient acceptability, low adherence due to discomfort, and electrodes that cause skin irritation in most people. Non-invasive wearable devices with automated photoplethysmography (PPG) acquisition could provide a convenient solution for accurate AF detection^3,4^. However, previous studies focused on short duration pulse oximetry data^5,6^ recorded in clinical environments^5–8^, and accounted for neither the significant motion artifacts to be expected in real-world environments nor the inclusion of premature atrial and ventricular contractions (PAC/PVC), which can degrade the accuracy of AF detection.

While it is relatively easy to detect PAC/PVC in ECG signals^9,10^, it is rather difficult to detect these rhythms in PPG as they do not provide a distinct waveform morphology from the normal sinus rhythm^11^. Another challenge with PPG for arrhythmia detection is that motion noise artifacts are a significant issue in smartwatch PPG data, as they can distort the PPG waveforms and mimic irregular dynamics seen in AF and PAC/PVC^4^, thereby, degrading the accuracy of arrhythmia detection.

Addressing the above-noted challenges requires large, diverse datasets collected in real life for long durations using smartwatches. However, long duration recordings of smartwatch PPG data require time-consuming adjudication of AF and PAC/PVC rhythms, aided by simultaneous recordings of ECG signals as the reference.

In this study, we addressed the above issues by using a large real-world smartwatch PPG dataset collected from our NIH-funded “Pulsewatch” clinical trial^12^. Two major novelties of our work are: (1) use of multimodal time-series data (PPG signal plus PPG-derived heart rates (HR), and accelerometer signal) combined with a simple 1D-Bi-GRU (bidirectional gated recurrent unit) network architecture that is computationally efficient for real-time assessment of multiclass cardiac arrhythmia detection; and (2) validation of the model on diverse PPG datasets from lab-controlled and real-life environments, thereby fully accounting for the effects of motion artifacts, long-duration recording, accuracy of PAC/PVC detection, and independent testing of the model to address the important issue of the generalizability of the model.

We employed a two-fold cross-validation approach, ensuring robust model evaluation by conducting large subject-independent testing on our Pulsewatch dataset. This method allowed us to assess the model’s performance across unseen participants, enhancing the generalizability of our findings. We also tested the generalizability of our models using two external datasets—the University of Massachusetts Medical Center (UMMC) Simband dataset, and the MIMIC III dataset—without using any of these datasets for training the model.

## Results

### Multiclass arrhythmia classification results on real-world smartwatch PPG

Fig. 1 shows two different multimodal input data configurations using three different databases along with our 1D-Bi-GRU network architecture. Fig. 2 shows that the Pulsewatch dataset was divided into three equally-sized subsets. Two of the subsets (which included AF, PAC/PVC, and NSR) served as the two-fold cross-validation and the last subset, which included only NSR subjects, was used for testing. The data folds were split to ensure independent subjects, thus, subjects in the testing dataset were not represented in the training dataset. Table 1 provides a detailed breakdown of the 166,904 segments collected from 106 subjects collected during the Pulsewatch trial, including the distribution of normal sinus rhythm (NSR), atrial fibrillation (AF), premature atrial contractions and premature ventricular contractions (PAC/PVC). These PPG segments were automatically detected as clean or relatively clean (if there was fewer than 5 seconds of motion noise artifact in a given 30-sec segment) by our previously developed motion artifact detection algorithms^13^ and the types of rhythms were adjudicated based on the rhythm shown in the reference ECG^14^.

**Table 1.** Subject and segment information for model development dataset (Pulsewatch) and independent testing datasets (Simband and MIMIC III).

| Datasets | Data segmentation | Num. of data Segments |  |  |  | Subjects |  |  |  |
| --- | --- | --- | --- | --- | --- | --- | --- | --- | --- |
|  |  | Total | NSR | AF | PAC/PVC | Total | NSR | AF | PAC/PVC |
| Pulsewatch<br>(training & testing data) | Total | 166,904 | 129,310 | 24,555 | 13,039 | 106 | 70 | 35 | 38 |
|  | Fold 1 | 58,318 | 39,356 | 12,265 | 6,697 | 36 | 18 | 17 | 19 |
|  | Fold 2 | 57,995 | 39,363 | 12,290 | 6,342 | 36 | 18 | 18 | 19 |
|  | Other NSR | 50,591 | 50,591 | 0 | 0 | 34 | 34 | 0 | 0 |
| UMMC Simband<br>(independent test data) | Total | 292 | 196 | 42 | 54 | 37 | 24 | 9 | 6 |
| MIMIC-III<br>(independent test data) | Total | 5,074 | 2,180 | 1,721 | 1,173 | 13 | 6 | 5 | 6 |

**Fig. 1.**
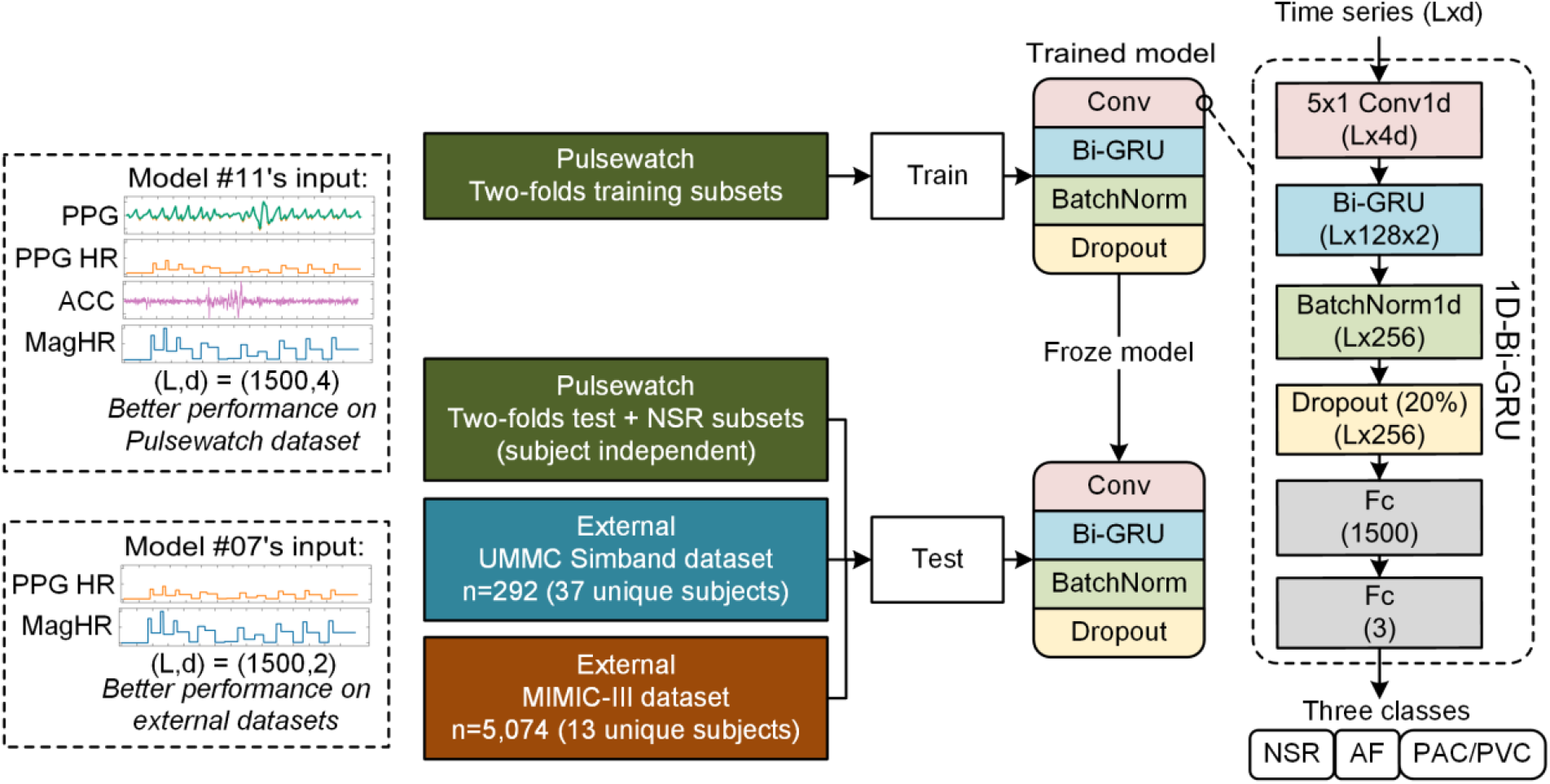
1D-Bi-GRU model’s input and architecture. The model with four inputs (PPG, PPG HR, ACC, and magnified HR) has the best subject-independent performance on the Pulsewatch dataset, and the model with only HRs (HR and magnified HR) has the best testing performance on external datasets that used different sensors and data collection locations than the Pulsewatch dataset.

**Figure 2.**
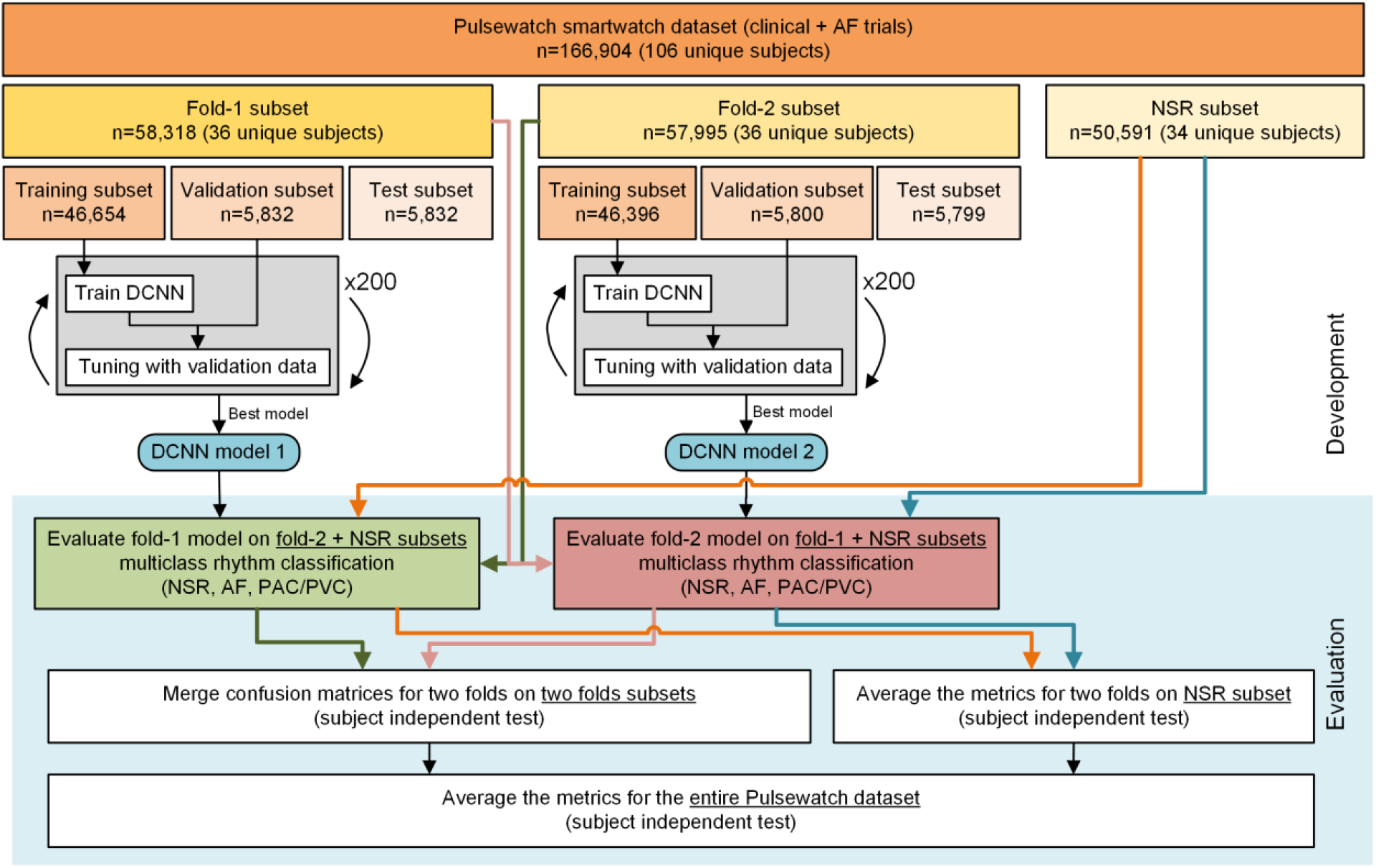
Workflow diagram for the subject-independent testing on Pulsewatch dataset.

Table 2 shows a comparison of our method, the 1D-Bi-GRU model, to two state-of-the art methods. We implemented two other state-of-the-art deep learning models^5,6^ and trained them using the same two fold cross-validation strategy on the Pulsewatch data so that we can directly compare these methods against our 1D-Bi-GRU model. Our method was also compared using various input signals, as noted in Table 2, to examine which combinations of the input signal provided the best arrhythmia classification results. Results of these comparisons using the Pulsewatch data are shown in Table 3. The best model is achieved by using all four modalities of input (model #12 using PPG, HR, ACC, and magnified HR, shown in Table 3). It showed the highest ever reported sensitivity of PAC/PVC detection at 83.52%. The previously highest reported PAC/PVC sensitivity on PPG data was 75.4%^6^ by the 1D-VGG-16 model. However, this result is based on the use of a fingertip PPG, which has a higher signal-to-noise ratio (SNR) than the smartwatch PPG we used, and the study was conducted to minimize motion artifacts. For fair comparison, we retrained the 1D-VGG-16 model^6^ with two types of input: (1) PPG waveforms as the sole input, and (2) the same four input signals (PPG, HR, ACC, and magnified HR (magHR)) from our Pulsewatch dataset that gave us the best performance for our deep learning model. When using only the PPG waveform as the input data, the 1D-VGG-16 model achieved a sensitivity of only 63.34% for PAC/PVC detection and an AF detection accuracy of 94.76%. These values are 20.18% and 2.55% lower, respectively, than those of our best model. With all four input signals, the 1D-VGG-16 model provided similar performance metrics for NSR and AF classification compared to our best model with the same four input signals. However, the sensitivity of PAC/PVC detection was only 71.32%, which is still lower than our result (83.52%) and their previously reported value of 75.4% with higher SNR data.

**Table 2.**
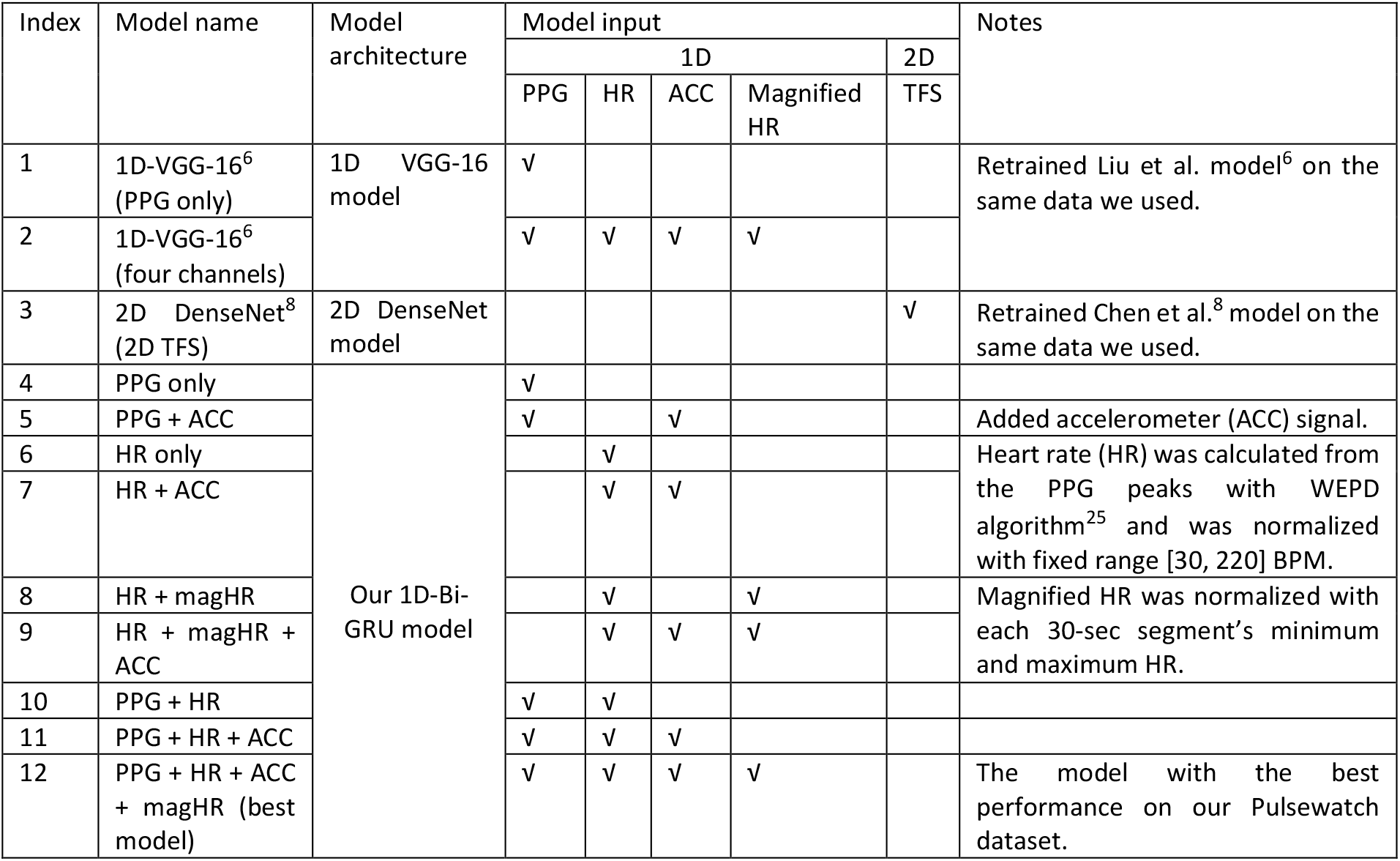
Input time series used by different models during the model development for multiclass arrhythmia classification.

| Index | Model name | Model architecture | Model input |  |  |  |  | Notes |
| --- | --- | --- | --- | --- | --- | --- | --- | --- |
|  |  |  | 1D |  |  |  | 2D |  |
|  |  |  | PPG | HR | ACC | Magnified HR | TFS |  |
| 1 | 1D-VGG-16<sup>6</sup><br>(PPG only) | 1D VGG-16 model | √ |  |  |  |  | Retrained Liu et al. model<sup>6</sup> on the same data we used. |
| 2 | 1D-VGG-16<sup>6</sup><br>(four channels) |  | √ | √ | √ | √ |  |  |
| 3 | 2D DenseNet<sup>8</sup><br>(2D TFS) | 2D DenseNet model |  |  |  |  | √ | Retrained Chen et al.<sup>8</sup> model on the same data we used. |
| 4 | PPG only | Our 1D-Bi-GRU model | √ |  |  |  |  |  |
| 5 | PPG + ACC |  | √ |  | √ |  |  | Added accelerometer (ACC) signal. |
| 6 | HR only |  |  | √ |  |  |  | Heart rate (HR) was calculated from the PPG peaks with WEPD algorithm<sup>25</sup> and was normalized with fixed range [30, 220] BPM. |
| 7 | HR + ACC |  |  | √ | √ |  |  |  |
| 8 | HR + magHR |  |  | √ |  | √ |  | Magnified HR was normalized with each 30-sec segment's minimum and maximum HR. |
| 9 | HR + magHR + ACC |  |  | √ | √ | √ |  |  |
| 10 | PPG + HR |  | √ | √ |  |  |  |  |
| 11 | PPG + HR + ACC |  | √ | √ | √ |  |  |  |
| 12 | PPG + HR + ACC + magHR (best model) |  | √ | √ | √ | √ |  | The model with the best performance on our Pulsewatch dataset. |

**Table 3.**
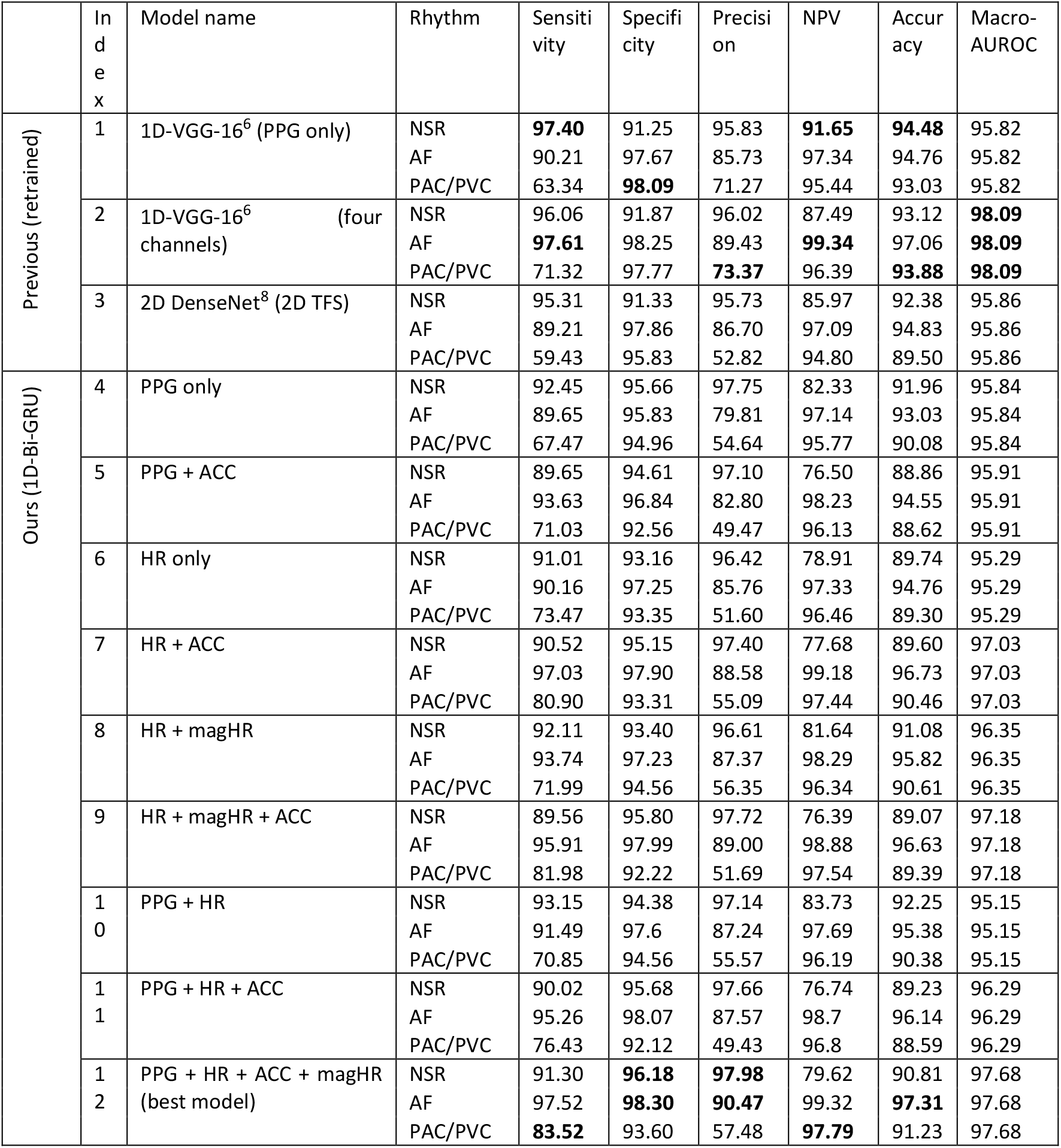
Subject-independent testing results for the multiclass classification of all models on the Pulsewatch dataset.

As most previous works^11,15,16^ only reported binary AF/non-AF classification results on PPG data, we merged the PAC/PVC class into the NSR class. We provide the performance metrics on binary AF classification in Table S2 in supplementary material. Our best model, as well as 1D-VGG-16^6^, have the highest binary AF classification results of >97% accuracy and >99% macro-averaged area under the receiver operating curve (AUROC). The two best models (ours and 1D VGG-16^6^) also achieved the highest macro-averaged AUROC of 99%, as shown in Table S2 in the supplementary materials. However, it should be noted that our model, 1D-Bi-GRU, has 93% fewer network parameters and is 3 times more computationally efficient than the 1D-VGG-16 model^6^ is, as detailed in Table 4.

**Table 4.**
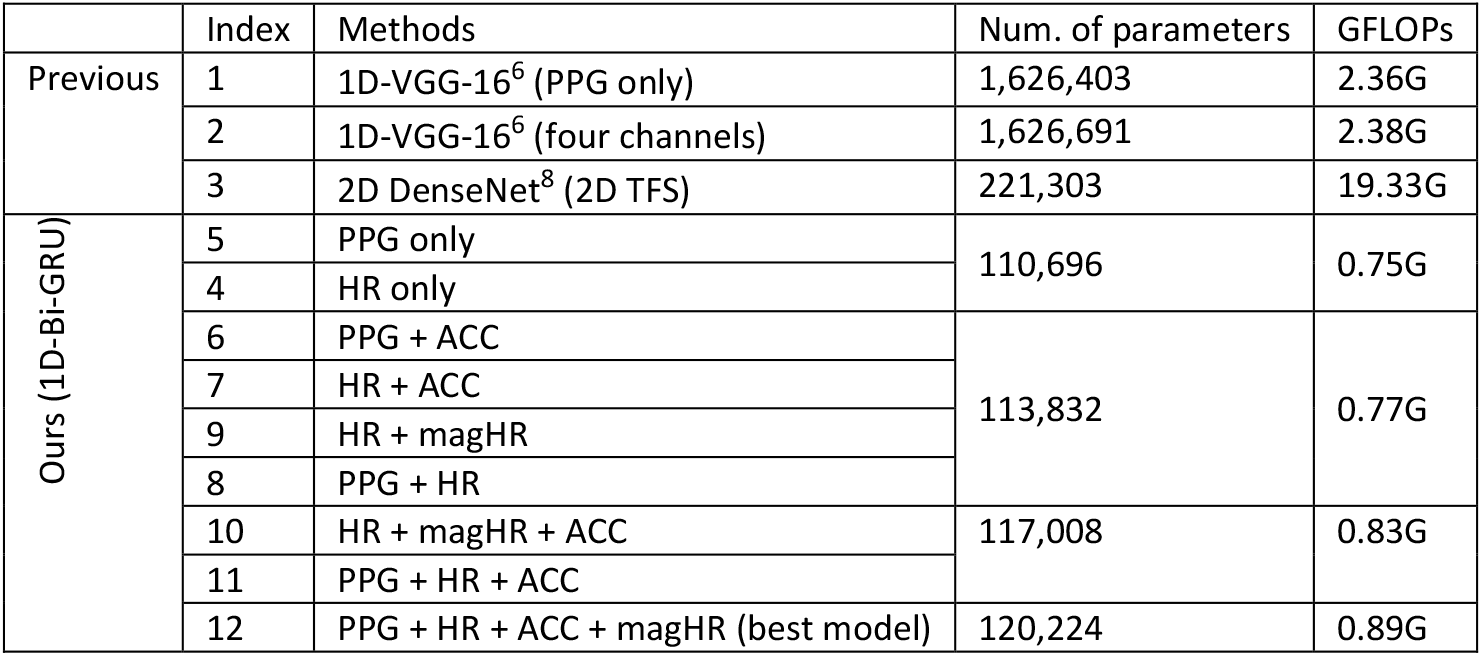
Computational Costs of all models.

### Improvement of multiclass arrhythmia classification using multimodal input

Table 3 shows the importance of including heart rate (HR) as one of the input signals, especially for increasing the sensitivity of PAC/PVC detection. With only HR (model #6 in Table 3), most of the multiclass classification results were comparable to those achieved by using only PPG signals as the input (model #4 in Table 3). However, the AF classification accuracy at 94.76%, is higher than the 93.03% AF classification accuracy with the PPG-only model. In addition, the sensitivity of PAC/PVC detection is 6% greater with HR only versus the PPG-only model. This highlights the importance of including heart rate derived features for arrhythmia detection. When both HR and PPG signals are included (model #10 in Table 3), the accuracy of NSR, AF, and PAC/PVC classification are 92.25%, 95.38%, and 90.38%, respectively, which are 0.29%, 2.35%, and 0.3% higher than they are for the model using only PPG (model #4 in Table 3), respectively.

In addition to HR, when adding the accelerometer (ACC) signal to the input, the multiclass classification performance metrics further improved. Comparing models #5 to #4, models #7 to #6, models #9 to #8, and models #11 to #10 in Table 3, by adding ACC to the input signals, the sensitivity values of AF and PAC/PVC increased on an average of 4% and 6%, respectively. This suggests the value of adding additional accelerometer information to further differentiate whether the change in PPG waveforms and the variations in HR were due to motion artifacts or cardiac arrhythmia. For example, in Fig. 3 (c), without the accelerometer information, the network model would have difficulty in knowing that the HR variations at around 24 to 26 seconds were caused by motion artifacts and were not due to premature beats. The notable amplitude changes in the ACC signal informed the network model to acknowledge that the corresponding PPG data are due to motion artifacts, hence, to override any dynamics that it might have otherwise concluded were reflective of either AF or PAC/PVC.

**Figure 3.**
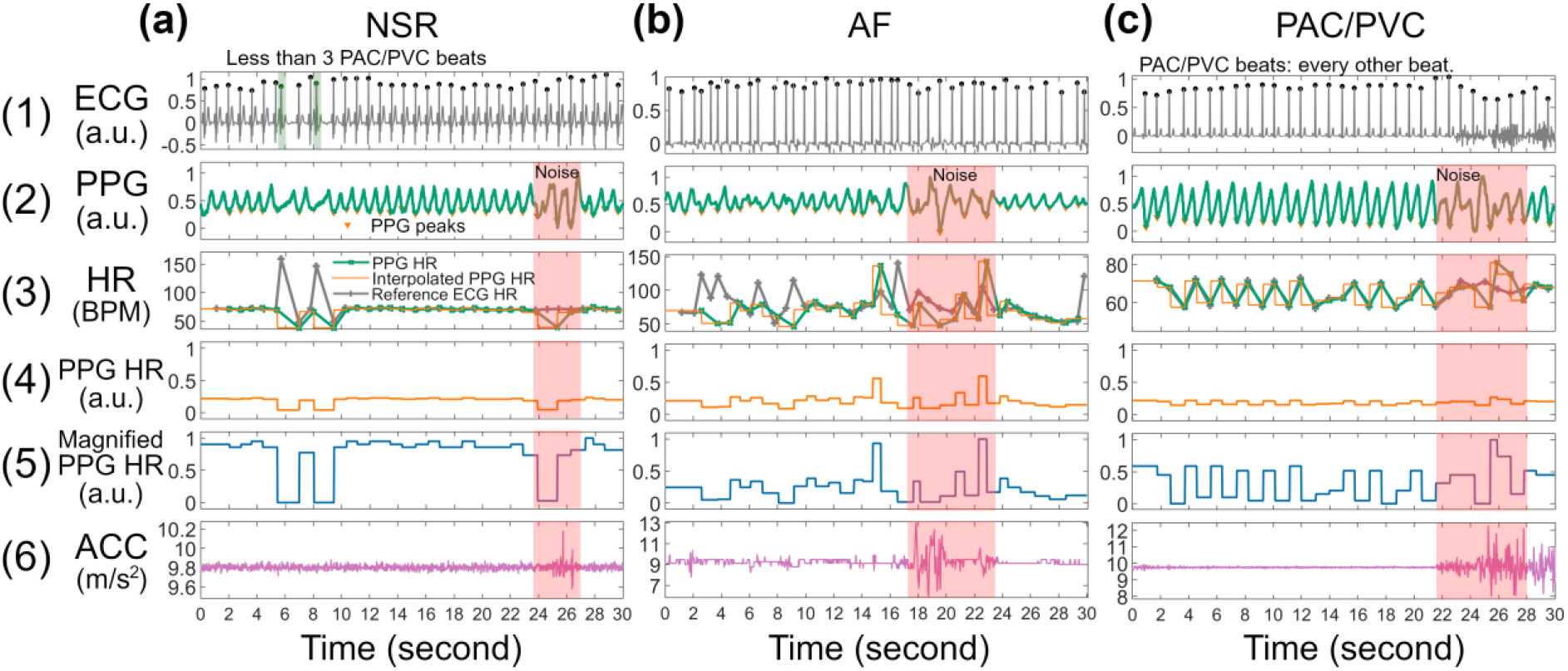
Example signal traces from (a) NSR, (b) AF, and (c) PAC/PVC with noisy section (≤5 sec of noise) highlighted in red. The (a) NSR segment contains less than three PAC/PVC beats. The (c) PAC/PVC segment contains bigeminy PAC beats (every other beat), therefore, the HR in the panel (3) has a zig-zag shape.

Lastly, we highlight the importance of proper normalization of HR dynamic range. The overall HR range (30-220 BPM) normalization reflects the mean HR in a 30-sec segment, but this approach depresses the local variations in HR, for example, seen in seconds 5-10 in panel 5 of Fig. 3 (a). Thus, we examined comparison of the overall versus local HR in arrhythmia classification performance. As shown in Table 3, we observe better overall performance with a local heart rate approach which we call magnified HR (magHR). The most notable improvement is the sensitivity of PAC/PVC detection with the use of magHR. For example, comparing models #12 to #11, with the input of magHR, the sensitivity of PAC/PVC improved from 76.43% to 83.52%. The magnified HR input provides to the deep learning model important details of local HR variations, such as the zig-zag shape visualized in Fig. 3 (c). This level of detail regarding a large and sudden change in HR (e.g., 15-20 BPM) is lost in the fixed range normalization, as it compresses the HR to nearly a flat line, as shown in panel 4 of Fig. 3 (c).

### Generalizability on external testing datasets

Two external independent datasets, the UMMC Simband dataset and MIMIC-III dataset, were used in this paper to illustrate the generalizability of our models. In other words, these two datasets were not used to train but they are solely used to test the network models. Table 1 shows the details of the UMMC Simband dataset and MIMIC-III dataset. Both datasets used different sensors than the smartwatch used in our Pulsewatch dataset, and the pulse oximetry data in the MIMIC-III dataset were recorded from a fingertip instead of wrist. Therefore, the PPG waveform of MIMIC-III was distinctly different and has a greater signal-to-noise ratio than the PPG waveforms recorded from a smartwatch (Fig. 4).

**Figure 4.**
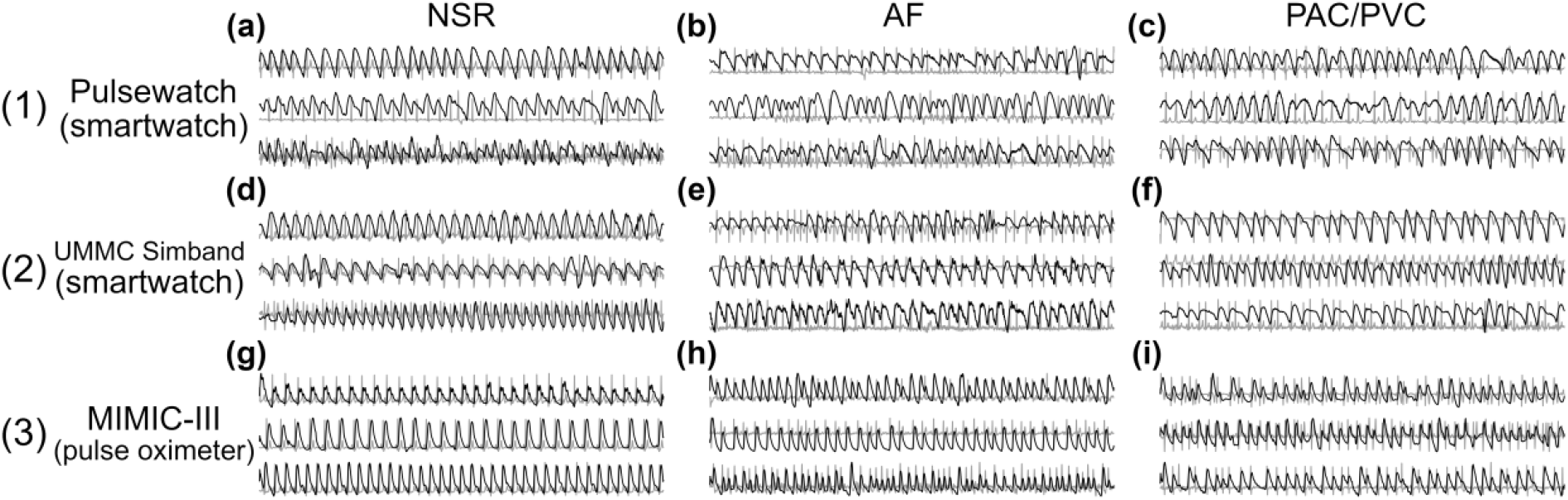
Example of PPG (foreground) and reference ECG (background) segments from (1) Pulsewatch, (2) UMMC Simband, and (3) MIMIC-III datasets.

Fig. 5 shows the macro-averaged AUROC of the Pulsewatch dataset (Fig. 5 (a)) along with subject-independent testing results from the UMMC Simband (Fig. 5 (b)) and MIMIC-III (Fig. 5 (c)) datasets. The best-performing model (#12 in Fig. 5 (a)) in the Pulsewatch dataset maintained superior performance on the UMMC Simband dataset (#12 in Fig. 5 (b)), demonstrating our network model’s robustness on data from untrained subjects. However, the model’s performance diminished slightly on the MIMIC-III dataset, likely due to the differences in the sensor modality. For the MIMIC-III dataset, the model using HR and magnified HR (#8 in Fig. 5 (c)) performed the best, and it also had consistent performance over Pulsewatch, UMMC Simband, and MIMIC-III datasets, with macro-averaged AUROC of 96.35%, 96.22%, and 94.17%, respectively (#8 in Table S3 in the supplementary materials). Our best model from the Pulsewatch dataset performed most accurately on the Simband dataset because the wrist PPG waveforms are similar in both datasets. However, this model’s performance deteriorated moderately on the MIMIC-III dataset, as the differences in the PPG waveforms in the development dataset and independent dataset were considerable (Fig. 4 row (3) for MIMIC-III vs. rows (1) and (2) for Pulsewatch and Simband, respectively). It is important to highlight that 1D-VGG-16 with four input data types—which was found to have only slightly less sensitivity on PAC/PVC detection when compared to our best model (#12) with the same number of input data types—had significantly smaller macro-averaged AUROC values for both Simband and MIMIC III databases, with the latter having only 87%. Note that for the Pulsewatch data, the macro-averaged AUROC value was 98% but this value decreased to less than 94% and 87% for Simband and MIMIC III datasets, respectively. However, our models maintained greater than 92% macro-averaged AUROC for all three datasets. Thus, our models show better generalizability than the 1D-VGG-16 approach does.

**Figure 5.**
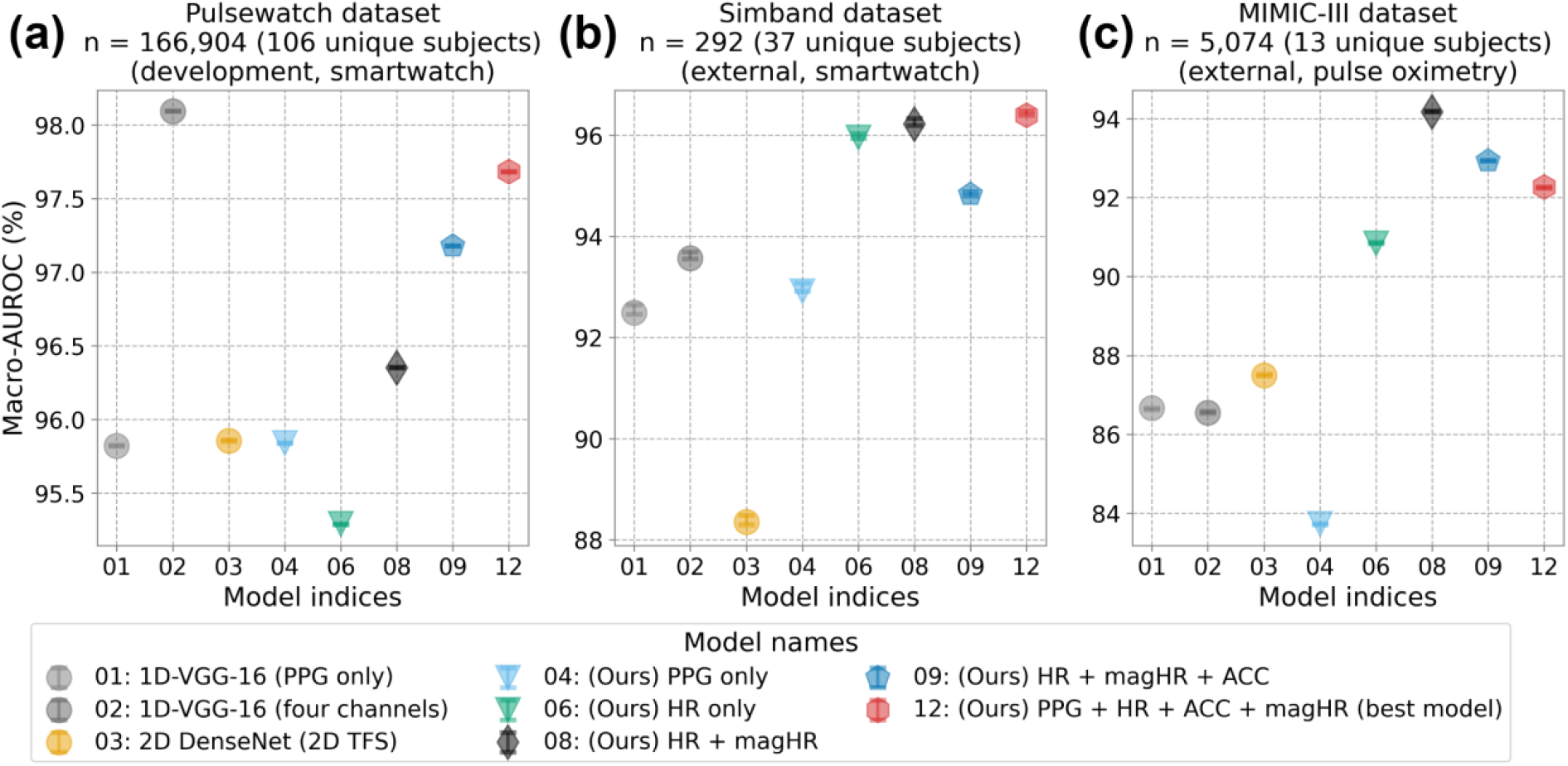
Macro-AUROC with 95% confidence interval of the subject independent testing results from (a) Pulsewatch, (b) Simband, and (c) MIMIC III.

### Computational efficiency of using multimodal input signal

In addition to achieving the highest sensitivity in PAC/PVC detection as well as the best performance metrics in AF classification, our multimodal 1D-Bi-GRU model has significantly fewer parameters (93% fewer) and has less computational cost (3 times less) than 1D VGG-16^6^. Table 4 shows the computational cost of all models. Our best model (#12) uses only 120,224 parameters, representing a 13.5-fold reduction in complexity compared to the 1D-VGG-16 structure^6^ (#1). While achieving superior performance, our model is more computationally efficient, enabling the potential for real-time processing on wearable devices. Our best model (#12) also requires only 0.89 Giga float-point operations per second (GFLOPs) to run a single instance, which is ∼3 times faster than the best comparison model^6^ (#1) and 21.7 times faster than 2D DenseNet model^8^ (#2) which uses 2D time-frequency spectrogram (TFS) as the input signal. Adding additional input signals to our model did not significantly increase the network’s parameters nor the computational costs.

## Discussion

### Real-world dataset using a smartwatch

This is the one of the first studies to perform multiclass cardiac arrhythmia classification on smartwatch PPG data collected for 14 days from stroke survivors’ daily life settings^12,17^. Our work differs from other prior work involving wearable PPG for the combined AF and PAC/PVC classifications, as they used fingertip PPG data^5,6^ and/or data were recorded in an in-hospital environment which minimized motion artifacts^5–7^. Adjudication is a labor-intensive task, and the labeling of AF/non-AF in one of the first publicly available smartwatch PPG datasets published along with the DeepBeat model^15^, may contain incorrect information. A study has found that while the DeepBeat model reported exceptional performance on the testing dataset (sensitivity: 0.98, specificity: 0.99, F1-score: 0.96), the performance was not satisfactory on the validation (sensitivity: 0.59, specificity: 0.995, F1 score: 0.69) and training datasets (sensitivity: 0.59, specificity: 0.998, F1 score: 0.74)^18^. Inaccurate ground truth labelling confuses any deep learning models, thereby degrading performance metrics when the models are confronted with non-trained testing datasets and other independent datasets generated from other studies^18^. Furthermore, the reference ECG data were not provided in DeepBeat, which precludes other studies, including our own, to benchmark against DeepBeat dataset’s performance metrics. In this study, we have painstakingly labelled 166,904 30-sec segments of Pulsewatch PPG data along with the corresponding reference ECG data, which we will provide to the public so that other investigators can use it to develop and benchmark their own algorithms’ performance. Another salient feature of our Pulsewatch dataset is that, because it was collected in a real-life environment over 14 days, the smartwatch PPG data contain diverse motion artifacts and provide many different characteristics of PPG waveforms that are representative of pAF and PAC/PVC. Note that there are not many databases with well-labelled PAC/PVC waveforms that are publicly available. Hence, our Pulsewatch along with Simband and labelled MIMIC-III PPG datasets will provide ample training data for other investigators working on this research topic.

### Multiclass classification on real-world smartwatch data

Despite the presence of challenging low-to-moderate motion corruption in the smartwatch PPG data (up to 5 seconds of noise per segment), our computationally efficient deep learning model achieved an unprecedented 83.52% sensitivity for PAC/PVC classification while maintaining high 97.31% accuracy for AF detection. Our study differs from previous work^11,15,16^, as their focus was on binary AF classification, while we performed a more challenging three-class classification for PAC/PVC, AF, and NSR. This approach provides better granularity for arrhythmia classification and may also lead to better AF detection. Although frequent PAC/PVC episodes have not been proven to directly cause AF^19^, an increased number of PAC/PVC episodes has shown positive correlation with a higher AF risk^20^. Therefore, accurately detecting PAC/PVC events provides finer details regarding cardiac arrhythmias. In addition, many previous studies have shown that frequent occurrences of PAC/PVC were one of the best features that can be used to predict AF using machine and deep learning approaches^20–23^.

### Heart rate as input to assist the training process

Another important finding is that using multimodal input signals, such as including HR calculated from the PPG as well as simultaneously recorded ACC, greatly increased deep learning model performance metrics compared to using only PPG. A previous study^24^ showed that a single-layer LSTM model with a sequence of 35 consecutive heartbeat periods resulted in inferior AF detection performance compared to a convolutional-recurrent neural network that used 30 seconds of PPG waveform data. In contrast, our HR-only model had compatible AF classification performance compared to our PPG-only model (see results of models #6 and #4 in Table 3). The HR information we used was not a simple linear interpolation between two consecutive heartbeats, but the heart rates were represented with a rectangular interpolation to better accentuate abrupt changes in heart rates. This transformation of HR is seen in panels 4 and 6 of Fig. 3. It should also be noted that the PPG peak detection algorithm we used to calculate HR was optimized to account for not only NSR but also AF and PAC/PVC, consequently, was proven to be one of the best algorithms for smartwatch PPG^25^, which resulted in the good performance of our model. This may explain why the previous study^24^ had a contradictory finding to ours, as most previous PPG peak detection algorithms were developed for NSR.

Although deep learning models using raw PPG waveforms as the input signal claim that they do not require extensive signal pre-processing and feature engineering^24^, these models often do require fine-tuning to account for the domain-shift problem when PPG signals are recorded from different anatomical sites^11^ and with different sensors. In contrast, our HR and magnified HR model (model #8) showed reliability and generalizability among different testing datasets, as shown in Fig. 5, suggesting that using HR instead of raw PPG waveforms may be a solution to address the domain-shift problem, especially as millions of raw PPG segments are not available to pretrain large deep learning models^26^. Furthermore, using HR as the input signal reduces the complexity of the network which further reduces computational costs, thereby enabling embedding of the algorithms into a smartwatch and other wearable form factors for real-time arrythmia detection.

### Accurate AF and PAC/PVC detection requires relatively clean PPG signals

Non-sudden motion-induced artifacts can introduce dynamic characteristics that are similar to AF, whereas sudden motion artifacts can mimic PAC/PVC patterns in PPG waveforms. In real-life recordings, fully clean PPG segments are difficult to obtain and if we only used completely clean segments, we would be left with only a small portion of the data. This is why the criterion for using a 30-sec segment was less than or equal to 5 seconds of artifacts in it. Consequently, allowing segments with a low-to-moderate amount of motion corruption explains the lower sensitivity of the NSR classification with our proposed models, as some segments were falsely classified as PAC/PVC. Hence, this issue is a trade-off between PAC/PVC sensitivity and NSR sensitivity. However, our approach, which resulted in 91% sensitivity for NSR, is sufficient since the primary aim is to better detect AF and PAC/PVC.

### Performance improvement with more PAC/PVC training data

It is our opinion that even greater than 83.5% sensitivity of PAC/PVC detection can be obtained with more training data for our models. The number of PAC/PVC events were far less than the number of AF events in our study, but this is typical. Hence, various data augmentation strategies including SMOTE and generalized adversarial network techniques as well as shuffling time locations where PAC/PVC occur in a given time series may lead to better generalized performance of any deep learning model. In addition, including a self-attention mechanism in a deep learning model to accentuate the occurrences of PAC/PVC waveforms may lead to further performance improvements.

## Methods

### Study population

#### Pulsewatch clinical and AF trials datasets

We recently completed a 2-year NIH-funded clinical trial named “Pulsewatch” (NCT03761394) to evaluate the accuracy of AF detection and usability of smartwatches for stroke survivors in real life conditions^12,17^. Participants who were randomized into the intervention group of Phase 1 (n=90) of the Pulsewatch clinical trial continuously wore the smartwatch system (which also included a smartphone for data collection) with a reference ECG chest patch for 14 days during their everyday lives. Detailed demographic and medical history information of the recruited participants (aged ≥50 with a history of ischemic stroke/TIA) can be found in ^12^.

As the Pulsewatch clinical trial progressed, only 11 participants (12%) were identified as having AF by the reference ECG patch out of the 90 subjects, and only 6 (6.7%) of them were confirmed as true AF subjects by cardiologists^12^. Although this ratio of AF is about the same as the 6.4% AF prevalence among the age group 65-69^27^, it would create a highly imbalanced dataset with a low number of AF subjects and segments for data analysis. Therefore, our co-authors at UMass Chan Medical School (UMCMS) conducted a separate AF trial simultaneously to enroll subjects with confirmed AF in clinic. For the first 30 enrolled participants, the experiment was conducted in-clinic, therefore, the recording duration was only about 20 minutes and did not provide the needed segments for balancing the AF class. For the later-enrolled 23 participants, the recording duration was extended to 7 days of free-living conditions to ensure enough AF segments would be recorded. The cut-off time of recording was 7 days because the battery of a single ECG chest patch lasted only 7 days.

The reference ECG was measured from the chest using a 2-lead rhythm patch device (Cardea SOLO, Cardiac Insight Inc., Bellevue, WA, USA) and wrist PPG data were collected using either Samsung Gear S3, or Galaxy Watch 3 (Samsung, San Jose, CA, USA). The patch ECG data, which were used as the reference for adjudicating PPG signals, consisted of one-channel ECG sampled at 250 Hz. The smartwatch data consisted of PPG signals and tri-axial accelerometer (ACC) signals which were converted to their magnitude values. The PPG sensor of the Samsung watch emitted a green light with a typical wavelength in the range of 520-535 nm. Smartwatch signals were all sampled at 50 Hz and were segmented into 30-sec lengths. The enrolled patients wore the ECG patch continuously and the smartwatch 23 hours a day with no restriction on their regular daily activities, for 14 consecutive days during the clinical trial. Due to the 7-day battery limitation, patients switched to a second new ECG patch on the 7th day of the trial. Smartwatches were charged daily for 1 h.

Formal ethical approval for this study was obtained from the University of Massachusetts Medical School Institutional Review Board (approval number H00016067 for the clinical trial and H00009953 for the AF trial). Written informed consent was collected from all patient participants.

#### Adjudication of Pulsewatch dataset

The adjudication of PPG segments was only performed on segments that were detected as clean and relatively clean (⩽5 seconds of motion noise) in the offline analysis using our previously developed motion artifact detection algorithms.^13^ This process was necessary since severe motion artifacts masked underlying arrhythmia information in PPG segments^28^. We also included those segments with ⩽ 5 sec of motion noise primarily to increase the number of usable PPG data segments for arrhythmia detection, as our previous study found that this amount of motion artifact did not result in many false positive AF alerts^4,13^.

The adjudication criteria for a 30-sec PPG segment for determining the types of rhythms were as follows:

1. AF segment: irregular rhythms (HR change ⩾ 10 BPM^3,29^ and missing P-waves in the reference ECG) must span the entire 30-sec segment.
2. PAC/PVC segment: needs to have three or more PAC/PVC beats^7^ in the reference ECG of a non-AF rhythm segment, and the definition of a single PAC/PVC beat is that it must have a heart rate (HR) change that is ⩾ 10 beats per minute (BPM)^7,25^ since an NSR beat typically does not vary more than 10 BPM^3,29^.
3. NSR segment: the remaining segments that were classified as neither AF nor PAC/PVC. It is possible that an NSR segment could contain one to two PAC/PVC beats.

Among the clean and relatively clean segments, the three types of arrhythmias were adjudicated in each 30-second segment by three experts^14^ using the aligned single-channel ECG as the reference. After applying our previously developed motion artifact detection algorithm followed by alignment of PPG and ECG signals, we found that 72 out of the 90 subjects in the clinical trial and 34 subjects from the AF trial had at least one clean/relatively clean PPG segment. Details of subject exclusion criteria are provided in Figs. S1 and S2 in the supplementary materials for the clinical trial and AF trial, respectively.

Table 5 lists the baseline characteristics of the population in the Pulsewatch clinical trial and Pulsewatch AF trial. It is clear that AF trial participants had higher AF burden (52.28%) among the confirmed AF subjects (AF burden was provided by the Cardea SOLO AF detection algorithm that was approved by the Food and Drug Administration (FDA)). The participants from the Pulsewatch clinical trial had a lower ratio of clean PPG segments. The average burden of PAC/PVC rhythm in the clean PPG segments among the participants in the Pulsewatch clinical trial was 11.60%, much higher than the 0.24% in the AF trial. The mean HR of each clean PPG segment from the AF subjects in the Pulsewatch clinical trial was 82.57 BPM, which is 5 BPM faster than the AF subjects’ mean HR in the AF trial. The mean HR of the NSR and PAC/PVC segments was similar in both trials, ranging from 67 to 69 BPM.

**Table 5.** Baseline characteristics of the Pulsewatch clinical trial and AF trial participants.

|  |  | Clinical Trial | AF Trial |
| --- | --- | --- | --- |
| Age (years) (SD) | | 65.33 ($\pm 9.08$) | 71.86 ($\pm 6.69$) |
| Female (%) |  | 41.67% | 38.10% |
| Race, non-white (%) |  | 11.67% | 9.52% |
| Mean ECG AF burden per subject (%) (SD) | Confirmed AF subjects, burden reported by Cardea Solo ECG | 3.60% ($\pm 1.55\%$) | 52.28% ($\pm 22.40\%$) |
| Mean clean PPG ratio per subject (%) (SD) | | 16.17% ($\pm 13.10\%$) | 31.71% ($\pm 12.69\%$) |
| Mean PPG rhythm burden per subject on clean PPG (%) (SD) | NSR | 86.72% ($\pm 30.60\%$) | 8.19% ($\pm 23.67\%$) |
| | AF | 1.68% ($\pm 12.80\%$) | 91.57% ($\pm 24.40\%$) |
| | PAC/PVC | 11.60% ($\pm 28.46\%$) | 0.24% ($\pm 0.74\%$) |
| Mean ECG heart rate on clean PPG (BPM) (SD) | NSR | 69.59 ($\pm 13.79$) | 62.00 ($\pm 17.75$) |
| | AF | 82.57 ($\pm 7.29$) | 77.40 ($\pm 12.02$) |
| | PAC/PVC | 67.84 ($\pm 7.64$) | 77.77 ($\pm 12.32$) |

#### Training and testing data segmentation for Pulsewatch dataset

The number of segments with AF and PAC/PVC differed widely among subjects, with some having many instances of these rhythms while others had few to none (Table S4 in the supplementary materials). Hence, how to determine which subjects to use for training and which for subject-independent testing became a challenging issue for addressing the generalizability of the algorithms. Therefore, given the imbalanced datasets (greater number of NSR than either AF or PAC/PVC), we used a two-fold cross-validation (CV) strategy as shown in Fig. 2, which divided the Pulsewatch dataset into two equal halves with the same number of AF and PAC/PVC subjects in both folds. Since the two folds (each fold represented by 36 unique subjects) also included many NSR segments (3 and 5 times more than the AF and PAC/PVC segments as shown in Table 2), including more NSR subjects would only make the training data more imbalanced. Therefore, the other 34 unique NSR subjects were only used for testing the algorithms. Table 1 shows that each fold has nearly the same number of subjects and data segments for all arrhythmia classes.

For each fold, we performed subject-based stratified random sampling to divide the data into 80% training, 10% validation, and 10% subject-dependent testing, as shown in the workflow diagram in Fig. 2. To evaluate subject-independent test results, the data from the first fold were used to test the second fold’s trained network model, and vice versa. The confusion matrices from the two folds were combined, and the arrhythmia classification metrics were calculated from this merged confusion matrix. These metrics were then averaged with those from the NSR-only test subset. This approach allowed us to report subject-independent testing results across the entire Pulsewatch dataset.

#### Independent testing database I: UMMC Simband dataset

This database along with MIMIC-III is from a study which performed three-class arrhythmia detection on smartwatch PPG data using statistical signal processing approaches^7^. Since both datasets were carefully adjudicated with a reference ECG, we used both datasets for the purpose of subject-independent testing. The UMMC Simband dataset was recorded from Simband 2 smartwatches (Samsung Digital Health, San Jose, CA, USA (henceforth referred to simply as Simband)), a different smartwatch than other commercially available smartwatches that were used in our Pulsewatch clinical trial, and data were collected in-clinic for 14 minutes^7,25^. While this dataset contains only 37 subjects with 292 clean segments, the dataset is the first smartwatch PPG dataset that is publicly available and labeled with three types of arrhythmias. Details regarding the number of subjects and the segments associated with each of the three types of arrhythmias are listed in Table 1. The age group of the subjects was the same as Pulsewatch (≥50 years of age), and detailed demographic information about this dataset can be found in reference^30^.

Both PPG signals and magnitudes of ACC signals from the UMMC Simband dataset were downsampled from 128 Hz to 50 Hz to match the Pulsewatch dataset’s sampling frequency. Only the green PPG channel was used for data analysis so that we are consistent with the green LED used for the Pulsewatch dataset^7^.

#### Independent testing database II: MIMIC-III dataset

We also used MIMIC-III’s PPG database as the second independent-subject test set to further evaluate algorithms. While ICU recordings for each subject in the MIMIC-III^31^ dataset contained hundreds of hours of data, we only used the subjects whose data had already been pre-processed and adjudicated for the AF study^32^. This subset of MIMIC-III consisted of 13 patients with 5,074 ECG segments and corresponding PPG segments. Details of the numbers of the selected subjects are listed in Table S5 in the supplementary materials.

Both ECG and PPG signals were segmented into 30-s lengths with no overlap. The ECG was used for PPG rhythm adjudication for each 30-sec segment. All signals were originally sampled at 125 Hz, but PPG signals were down-sampled to 50 Hz to be concordant with the Pulsewatch dataset.

### Signal Preprocessing of the Time-Series Data

#### 1D Time Series Data Preparation

The left, middle, and right top rows of Fig. 3 show representative ECG signals for normal sinus rhythm (NSR), AF, and PAC/PVC, respectively. Row 2 of Fig. 1 shows the corresponding and simultaneously measured PPG, filtered with a 6th-order Butterworth bandpass infinite impulse response (IIR) filter (0.5 to 20 Hz)^25^ to remove baseline wandering as well as high frequency noise. Each filtered PPG was then normalized to [0, 1] based on each segment’s minimum and maximum values, ensuring uniform scaling for subsequent processing. The third row shows HRs obtained via ECG and the corresponding PPG along with interpolated PPG HR (shown in orange lines), which better captures abrupt HR changes than simply connecting two consecutive HR points. The interpolation method used for extracting PPG HR are further described in the next section.

We included HR as an input to the deep learning models, which has several advantages even when compared to using millions of PPG waveforms as the sole input^26^. Our prior work has shown that cardiac arrhythmias can be accurately discriminated using HR^3^. As shown in Fig. 3, PPG waveform distortions seen especially for PAC/PVC and AF are better captured with changes in HR. We also included ACC as an input signal to the network models so that they can be trained to discriminate between true arrhythmia (e.g., AF and PAC/PVC) versus motion artifact induced “arrhythmia”.

The fourth and fifth rows of Fig. 3 show normalized PPG heart rates and magnified PPG heart rates, respectively, where the fourth row was normalized within a [30, 220] BPM range to represent those with rapid ventricular response (RVR) (e.g., heart rates >100 BPM)^25^, such as in Fig. 3 (b). The fifth row represents each segment’s minimum and maximum HR values so that non-RVR rhythms can be represented with better dynamic ranges, such as the sudden drop of the PPG HR in the NSR segment in Fig. 3 (a), and the zig-zag shaped HR in the PAC/PVC segment in Fig. 3 (b). The tri-axial accelerometers’ (ACC) magnitudes in the 0 to 20 m/s^2^ range (daily activity range) are shown in row 6 of Fig. 3.

Since the accelerometer data in Simband data is in the numeric value of gravitational acceleration (e.g., 1 G if Simband remains stationary), we converted ACC signals of Simband data into the unit of m/s^2^ by multiplying them by 9.8. Since MIMIC-III did not record any accelerometer data, we used a constant 9.8 m/s^2^ value for the ACC signal.

#### Extraction of PPG Heart Rates (HR)

The HR for each PPG segment was calculated using the waveform envelop peak detection (WEPD) algorithm, as this approach has been shown to be one of the most accurate and can account for various arrhythmias in PPG signals^25^.

#### Machine Learning Model Design: 1D-Bi-GRU Model

While most prior works used complicated and large structured deep learning models for multiclass arrhythmia classification using PPG signals, such as 1D-DenseNets^5^, 1D-VGG-16^6^, and 2D-DenseNets^8^, we illustrate in this work that a simple and computationally lightweight model using 1D bi-directional Gated Recurrent Unit (1D-bi-GRU)^34^ can reach similar classification performance. This time-series based model has shown its ability to detect motion artifacts from PPG^34^, and it is also particularly well-suited for cardiac arrhythmia classification. GRU learns the long-term dependencies in the time series better than does the recurrent neural networks (RNN), and it also has fewer parameters to capture the dynamics of the data and is computationally faster than the long short-term memory (LSTM) structure due to having only one hidden state, compared to two states in LSTM^35^. The update gate in the GRU replaces the complicated forget and input gates of LSTM^35^.

As described in ^34^ and shown in Fig. 1, our input time series has a dimension of (L, d) (L=1,500 samples in our case, while d is the number of input channels). The first layer is a 1D convolutional neural network (CNN) to embed the input time series with 4d filters with a kernel size of 5, a stride size of 1, and a padding size of 2, hence, the output dimension is (L, 4d). The second layer is a bi-GRU layer, which combines the outputs of two GRU networks (with 128 units each) that process the input-embedded information in opposite directions, allowing for each sample to consider both preceding and proceeding samples. A batch normalization is then applied, followed by a dropout of 20% to avoid overfitting. Lastly, a dense layer combines the output of the previous layers (L, 256) into a dimension of (L, 3) for predicting three classes (0=NSR, 1=AF, and 2=PAC/PVC).

#### Machine Learning Model Training Process

As shown in Table 1, the number of NSR segments is 3 and 5 times more than AF and PAC/PVC segments, respectively. To prevent over-fitting, up-sampling of the minority PAC/PVC and AF classes to the same number of segments in the majority NSR class was implemented in the training and validation sets to ensure unbiased performance in the testing data.

A batch size of 32 was used, as it showed a faster and more stable training validation process compared to a batch size of 512. The cross-entropy loss function was used for three-class classification. The Adam optimizer was used with the same parameters as described in reference^34^. We trained the models with a maximum of 200 epochs and selected the best model using the minimum validation loss. Early stop was used if the validation loss did not improve in 40 consecutive epochs.

#### Evaluation Metrics

For the performance evaluation of the proposed and other compared methods, we calculated five key metrics: sensitivity, specificity, precision, negative predictive value (NPV), and accuracy in keeping with other publications ^5–7^. The micro-averaged area under the receiver operating characteristic (AUROC) curve cannot reflect the problem of imbalanced dataset^5,6^, hence, we chose macro-averaged AUROC instead to give equal importance to each class. However, we still included the micro-averaged AUROC value in Tables S1, S2, and S3 for completeness and comparison to other works which reported this metric.

## Data Availability

The Pulsewatch data (our segment-level adjudication and continuous recording of reference ECG, PPG, and ACC) used in this paper will be publicly available for downloading on https://www.synapse.org/Synapse:syn23565056/. The external testing datasets?UMMC Simband dataset and the MIMIC-III dataset?are already publicly available.

https://github.com/Cassey2016

https://www.synapse.org/Synapse:syn23565056/

## Data availability

The Pulsewatch data (our segment-level adjudication and continuous recording of reference ECG, PPG, and ACC) used in this paper will be publicly available for downloading on

https://www.synapse.org/Synapse:syn23565056/. The external testing datasets—UMMC Simband dataset and the MIMIC-III dataset—are already publicly available^7^.

## Code availability

The code of our proposed 1D-Bi-GRU model and its trained version on different input time series, the code of the model training and evaluation on all three datasets, as well as our implemented version of the two comparison methods^6,8^ will be publicly available to download on https://github.com/Cassey2016. The code and documentation for loading Pulsewatch data will also be released.

## Acknowledgements

This work was supported by NIH under Grant R01 HL137734.

## Notes

### Competing Interest Statement

The authors have declared no competing interest.

### Clinical Trial

NCT03761394

### Clinical Protocols

https://doi.org/10.1016/j.cvdhj.2021.07.002

